# Association between population density and infection rate suggests the importance of social distancing and travel restriction in reducing the COVID-19 pandemic

**DOI:** 10.1101/2020.09.04.20187849

**Authors:** Heliang Yin, Tong Sun, Lan Yao, Yan Jiao, Li Ma, Lin Lin, J Carolyn Graff, Lotfi Aleya, Arnold Postlethwaite, Weikuan Gu, Hong Chen

## Abstract

**Background:** Currently, the 2019-nCoV has spread to most countries of the world. Understanding the environmental factors that affect the spread of the disease COVID-19 infection is critical to stop the spread of the disease. The purpose of this study is to investigate whether population density is associated with the infection rate of the COVID-19.

**Methods:** We collected data from official webpages of cities in China and in the US. The data were uploaded on Excel spreadsheets for statistical analyses. We calculated the morbidity and population density of cities and regions in these two countries. We then examined the relationship between the morbidity and other factors.

**Results:** Our analysis indicated that the population density in cities in Hubei province where the COVID-19 was severe was associated with a higher percentage of morbidity, with an r value of 0.62. Similarly, in the US, the density of 51 states and territories is also associated with morbidity from COVID-19 with an r value of 0.55. In contrast, as a control group, there is no association between the morbidity and population density in 33 other regions of China, where the COVID-19 epidemic is well under control. Interestingly, our study also indicated that these associations were not influenced by the first case of COVID-19. The rate of morbidity and the number of days from the first case in the US has no association, with an r value of -0.1288.

**Conclusions:** Population density is positively associated with the percentage of patients with COVID-19 infection in the population. Our data support the importance of such as social distancing and travel restriction in the prevention of COVID-19 spread.

## Introduction

At present, there is a worldwide pandemic of COVID-19 [Wang, Horby, Hayden, & Gao, 2020]. With its associated morbidity and mortality, COVID-19 is on the track to become one of the most catastrophic pandemics in human history [Wang et al., 2020]. A considerable amount of research and publications have focused on the analysis of the factors that lead to COVID-19 infection [Koo et al., 2020; Sun et al., 2020]. After its person-to-person transmission was confirmed, the influence of environmental factors on COVID-19 transmission in human populations has received considerable attention. Several environmental factors, such as the nature of surfaces of objects and the role of transmission airborne infections, have attracted great attention from the public. Further systematic understanding of the impact of environmental factors on human-to-human transmission of this virus may be extremely important in designing measures to contain the COVID-19 epidemic [Bedford et al., 2020; Jarvis et al., 2020; Lasry et al., 2020]. Among many environmental factors, population density is one of the conditions that cannot be underestimated and may affect the infection rate of COVID-19. Population density may directly reflect on whether and how social distancing and travel restriction work to slow the spread of COVID-19 [Gewin et al., 2020; Gibson et al., 2020]. In theory, the higher the population density, the greater the chance of COVID-19 infection. However, it is still unclear to what extent population density affects the infection rate of COVID-19.

The current available data in coronavirus infections provides an opportunity to conduct a preliminary analysis of the impact of population density on the infection rate of COVID-19. For example, the number of people infected in different cities with different population densities in China has been reported [Chen et al., 2020; Huang et al., 2020]. COVID-19 infection rates and mortality rates in different states and territories of the United States are currently reported and updated daily [Lasry et al., 2020]. Here we present a preliminary analysis of the impact of population density on the rate of COVID-19 infections.

## Method

### 1 Data collection from different regions and cities located in China and the US

Data collection were from three sets of multiple locations. The first set of locations is the 17 cities in the Hubei province in China. These cities are all located close to Wuhan and had large numbers of COVID-19 patients among cities in China. Data from these cities were from the official website of Hubei province. Next, we collected second set of data from other cities and provinces of China, hereafter referred to as other regions. The differences between these other regions and cities in Hubei is that less people contracted COVID-19 in other regions than in the cities of the Hubei province. COVID-19 disease in these other regions was well tracked, and disease incidence did not reach thepandemic level. The data from these other regions are all from provincial websites (http://www.hubei.gov.cn/fbjd/dtyw/). The third set of data is from the 50 states and three territories of the US. These data were obtained from a website (https://www.cdc.gov/coronavirus/2019-nCoV/index.html). Data collected include the names of the city/state/territory, accumulative deaths, population size in (millions) geographical area in (square kilometers).

### 2. Data organization and calculation

Data were uploaded and organized using Excel software. The population density of a city was calculated by dividing the population of the city by the geographical area. Thus, the result was the number of persons per square kilometer. The infection morbidity rate of a given location was obtained by dividing the number of COVID-19 cases by the population of the city. The relationship between the population density and the disease morbidity then was analyzed by correlation coefficients. Linear regression model was used to demonstrate their mathematic relationship.

### 3. First cases in the US and territories

In order to examine the relationship between the disease morbidity and the time of the first case in a city, we collected the information on the first cases in these cities and coded their first cases for the analysis. Based on dates of reported onsets of the first cases in the US states and territories, we assigned numbers based on the date of the first case. We first assigned the earliest US case in Washington State in Seattle on January 21, 2020 as #1. The number assigned to the other locations is derived by adding 1 to the difference between date of the first case in a given city and the day of first case in Seattle, January 21, 2020. For example, the first case reported in Arizona was on January 26, 2020. The Arizona number therefore is 1+(January 26 – January 21) = 6.

### 2 Statistical analyses

For the correlation, we used the previous criteria to categorize the strength of the correlation (Wang et al., 2016). Thus, if the r value is greater than or equal to 0.7 or is less than or equal to -0.7, it is a strong positive or negative correlation, respectively. If the r value is between 0.35 and 0.69 or -0.35 and -0.69, a correlation exists. However, r values falling between 0 and 0.35 or 0 and -0.35, were regarded as not correlated.

## Results

1. Basic patterns and information from different regions Our sets of data include cities of Hubei province, other regions in China, and 53 states and territories in US. The first set of data was obtained from 17 cities in the Hubei Province (Supplemental Table 1). These 17 cities had a total of 68,128 patients diagnosed with COVID-19 and 4,512 deaths from COVID-19. The total population in these 17 cities is 59.965 million, living in an area of 162,245 square kilometers. The calculated average COVID-19 mortality rate is 0.665%_0_. The second set of data is the population density and the total number of people in other provinces and cities or other regions in China (Supplemental Table 2). The collected data show that the average population density of these cities and provinces is 1,106 square kilometers. However, the morbidity rate in these places is very low, and the prevalence rate is 0.025 per thousand. The third set of data is the population density and prevalence of 53 major cities in states and territories in the United States (Supplemental Table 3). The population density of these cities is averaged to be 90 per square kilometer. The morbidity is 1.412 per thousand people.
2. Association between population density and mortality in Hubei province. Our data indicate that population density of the 17 cities in the Hubei province is positively associated with the disease mortality. There are considerable differences in the population density and death rate among these 17 cities. Although on average, the population density is 489 persons per square kilometers, the difference from city to city is large, ranging from 24.6 in Shennongjia to 1461 in Enshi (Figure 1A). Similarly, on average, the average disease morbidity is rate is 3.49% ranging from 0.025 to 4.167% (Figure 1B). Correlation analysis indicated that there is a positive association between the population density and disease morbidity, with an r value of 0.620 (Figure 1C).
3. Association between population density and morbidity in major cities in the US states and territories. Our data indicate that population density in the US states and territories is positively associated with disease morbidity. The population density of the US states and territories is much lower than that of China; nevertheless, there are also considerable differences in the density and death rate among different locations in the US states and territories. On average, the US population density is 80 persons per square kilometer; the difference from state to state is from as low as 0.49 in Alaska to as high as 438.00 in New Jersey (Figure 2A). Similarly, on average, the disease morbidity has a rate of 1.41%, with a variation from 0.07% in South Carolina to 9.94% in New York (Figure 2B). Further comparison indicates that there is a positive correlation between the population density and disease morbidity, with an r value of 0.552 (Figure 2C). There is apparently a highly consistent relationship between population density and COVID-19 mortality. Because the date of the first cases reported in different cities in the US are considerably different, and the disease epidemic is still ongoing, we analyzed the relation between the date of the first case and the morbidity to determine whether the date of the first case influenced the rate of morbidity (Supplemental Table 4). Surprisingly, there was no association between them, with an r value of -0.1288.
4. Non-association between population density and mortality among other regions in China Since the disease in other regions of China was well under control, an epidemic did not materialize into the general population before elimination of the infection source. We hypothesized that in this case the population density in other regions would not be associated with the disease morbidity. These regions include 33 regions, with an average of 1,106 persons per kilometer but with large differences from 2.80 persons per kilometer in XiZang to 13,984 persons per kilometer in Aomen (Figure 3A). The disease morbidity rate is low, all below 0.1%, except Shandong which had a rate of 0.4% (Figure 3B). The r value for the correlation between population density and disease morbidity rate is 0.04 (Figure 3C). Thus, the data from other regions in China services as a negative control; when the COVID-19 disease does not morph into an epidemic, population density is not associated with the disease morbidity.
5. Difference between China and the US on the measures of social distance and the impact of disease epidemic. By analyses of the data from the Hubei province in China and 53 major US cities in US states and territories, we obtained the positive correlations between the population density and the disease morbidity. We next asked whether there is a difference between these two data sets of cities on the impact of the disease epidemic. We calculated the morbidity of population density from 100 from 1000 using the linear formula obtained from the Hubei and major cities of US states and territories. Although both data sets have positive correlation between the population density and morbidity, they are not at the same degree. Based on the formula derived from Hubei province, the morbidity increased from 0.177 to 1.377 when the population density increased from the 100 to the 1000 (Figure 4A). On the other hand, in major cities and territories of US, the mortality increased from 1.453 to 7.93 when the population density increased from the 100 to the 1000 (Figure 4B). The increases of rates between these two sets of data are significantly different (Figure 4C).

**Figure 1.**
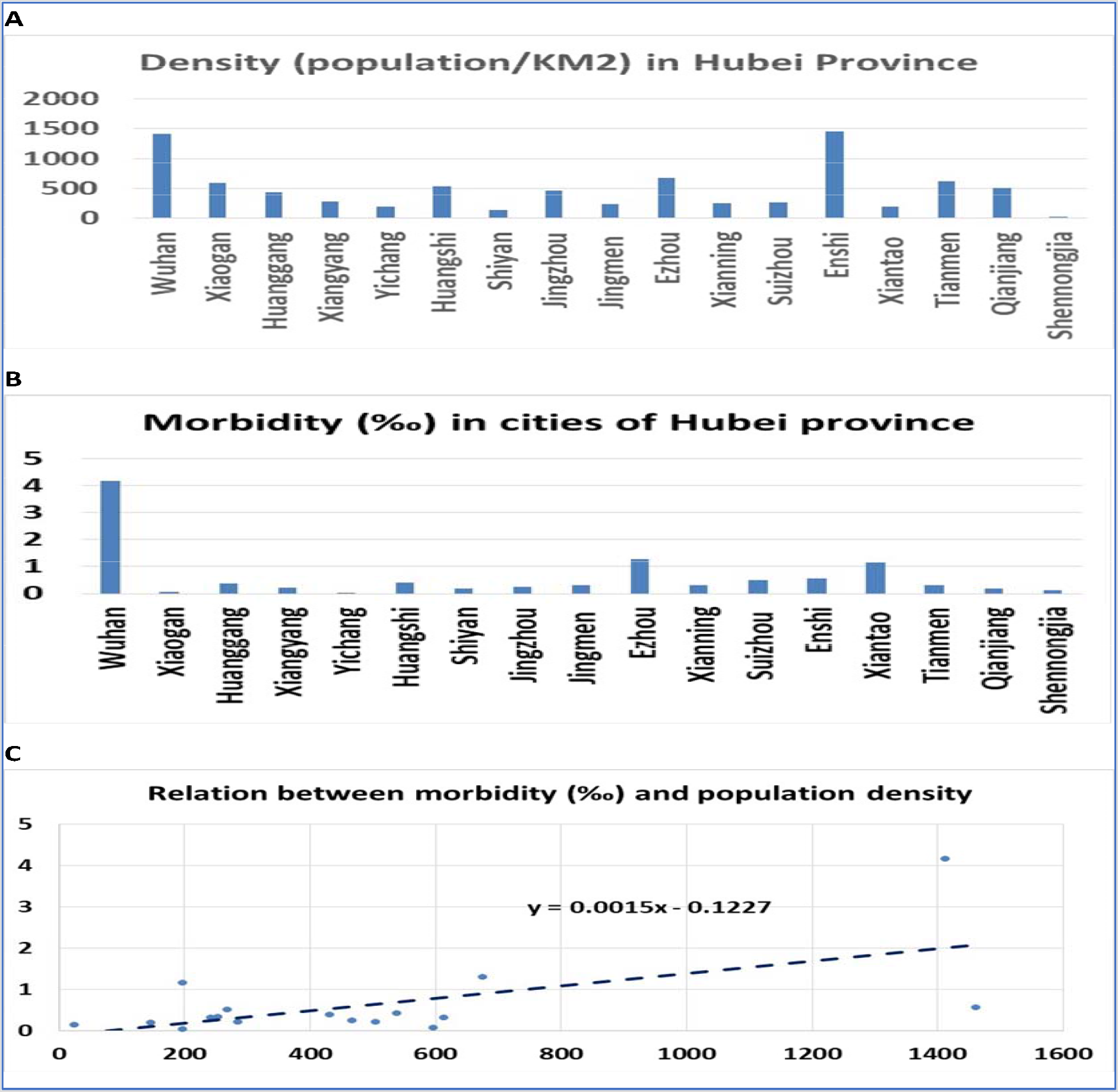
Population density and morbidity in Hubei province. Fig. 1A: Population density in the 17 cities in Hubei province. The number on the vertical axis is the average number of persons per kilometer. Names of the cities are listed on the horizontal axis. Fig. 1B: Disease morbidity rate in the 17 cities in Hubei province. The number on vertical axis is the average rate of morbidity. Names of the cities are listed on the horizontal axis. Fig. 1C. The relationship between the population density and disease morbidity. The number on vertical axis is the disease morbidity rate, and the horizontal axis is the number of persons per kilometer.

**Figure 2.**
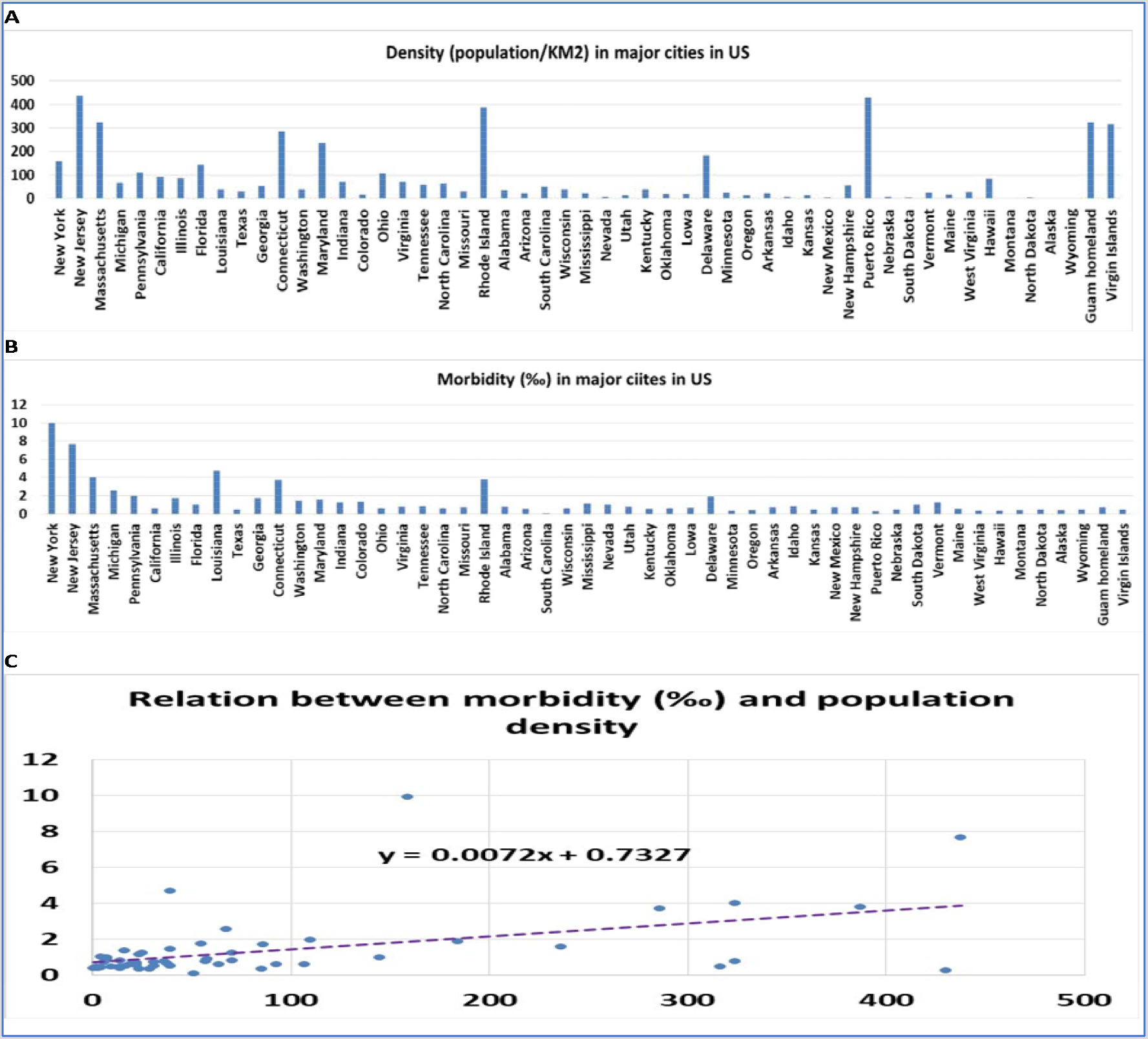
Population density and morbidity in 53 locations in the US and its territories. Fig. 2A: Population density in the 53 locations in US states and territories. The number on vertical axis is the average number of persons per kilometer. Names of the states and territories are listed on the horizontal axis. Fig. 2B: Disease morbidity rate in the 53 in US states and territories. The number on vertical axis is the average rate of morbidity from COVID-19. Names of the states and territories are listed on the horizontal axis. Fig. 2C. The relationship between the population density and disease morbidity. The number on vertical axis is the disease morbidity rate, and the horizontal axis is the number of persons per kilometer.

**Figure 3.**
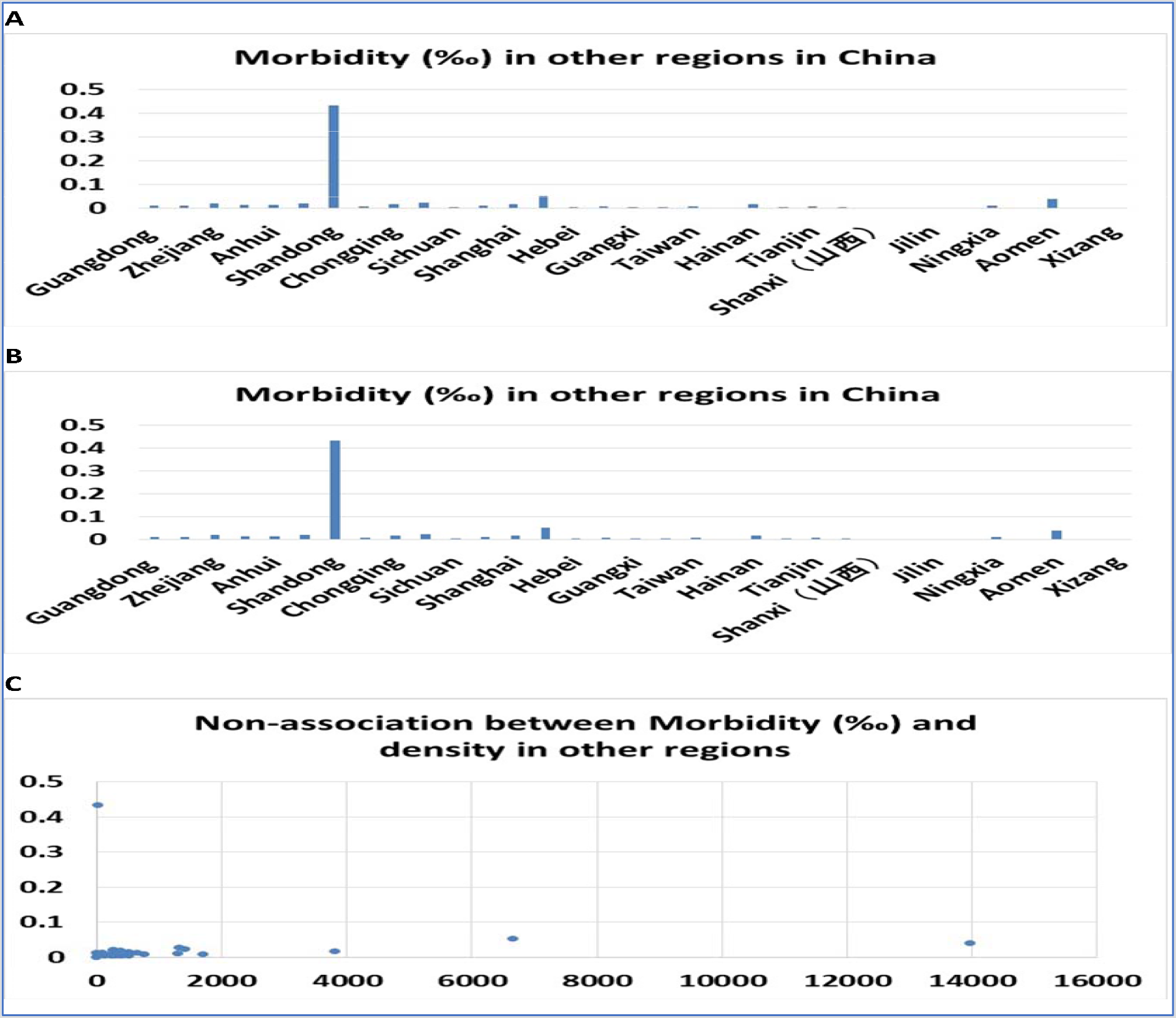
Non-association between population density and morbidity in other regions in China. Fig. 3A: Population density in the 33 regions in China. The number on vertical axis is the average number of persons per kilometer. Names of the cities are listed on the horizontal axis. Fig. 3B: Disease morbidity rate in the 33 regions in China. The number on the vertical axis is the average rate of morbidity. Names of the cities are listed on the horizontal axis. Fig. 3C. The relationship between the population density and disease morbidity. The number on vertical axis is the disease morbidity rate, and the horizontal axis is the number of persons per kilometer.

**Figure 4.**
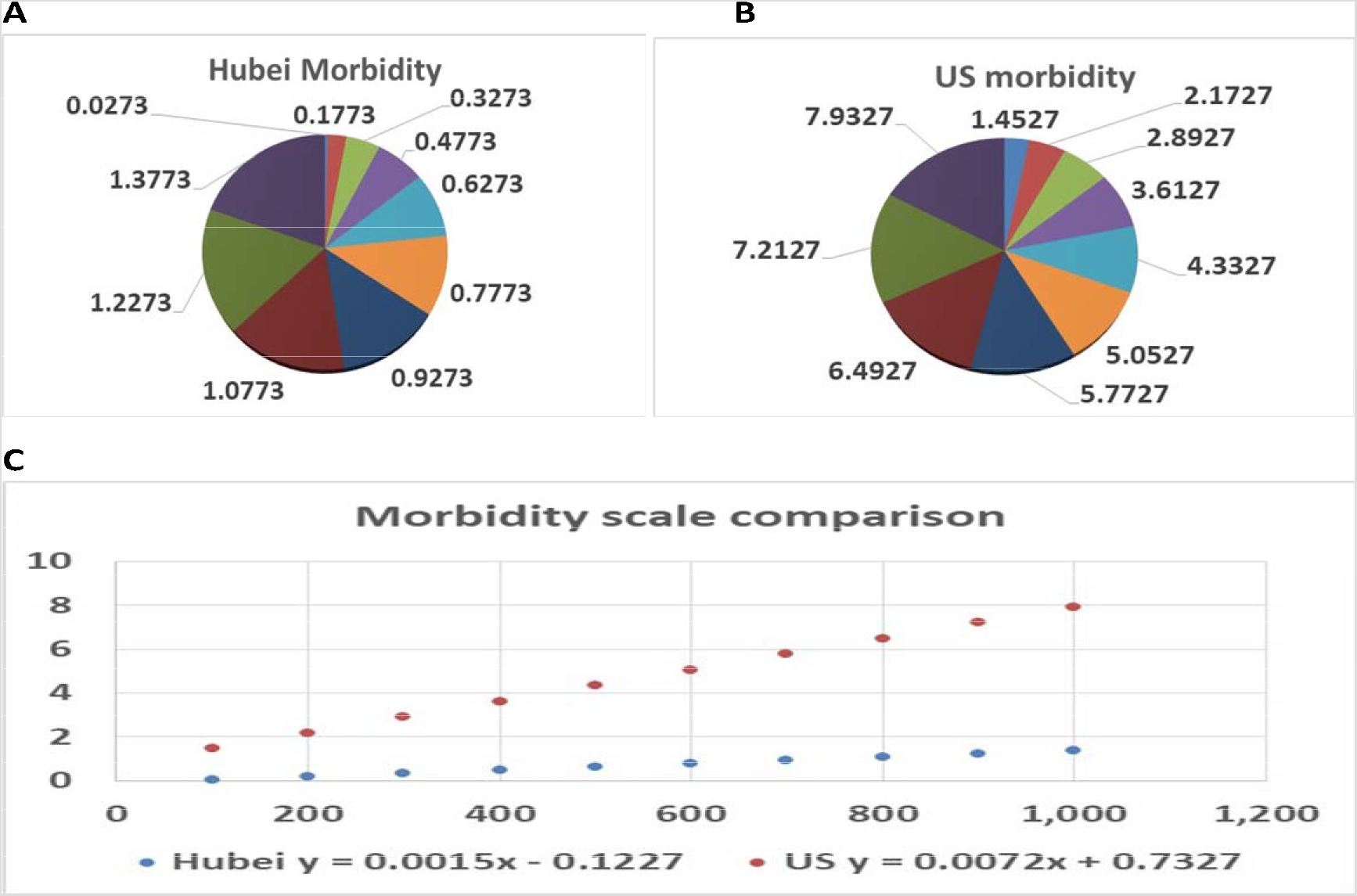
Comparison of the morbidity scales between major cities in the Hubei province of China and US states and territories. Fig. 4A: The morbidities at ten different scales calculated based on the formula derived from 17 cities of the Hubei province in China. Fig. 4B. The morbidities at ten different scales calculated based on the formula derived from 53 major cities of US states and territories. Fig. 4C. The comparison of disease morbidities between cities in China and in US states and territories when the population density increases from 100 to 1000 persons per kilometer. The number on vertical axis is the disease morbidity rate, and the horizontal axis is the number of persons per kilometer.

## Discussion

Our analyses clarified the positive correlation between population density and epidemic infection rate. From the data of 17 cities in the Hubei province, the correlation between population density and the number of people infected with COVID-19 in this epidemic is 0.62, showing a relatively high positive correlation. An analysis of 53 locations in US states and territories yielded a correlation of 0.55 when examining the relation between population density and the number of people previously infected. Additionally, the number of infected patients is not directly related to the date of initial infection. These two sets of data show that due to the higher population density, the chance of contact between people is relatively high. When the coefficient representing the relation between population density and number of persons infected is relatively high, the absolute number of people who contact each other is relatively high. The findings from this study indirectly support the practice of keeping social distance and enforcing of travel restriction.

Our data showed that given the same population density, the scale of disease morbidity in US states and territories is much higher than that in China. Given the complex differences between the social and economic systems and ethnic populations in these two countries, the reason for such a difference needs to be explored in a much broader investigation. Our study only reveals that such a difference exists but does not explain why there is such a difference.

In addition, this study also shows that in the case when the epidemic is well controlled, the initial source of infection is detected early and the route of infection could be cut off. In the control set, when there is no major outbreak, the population density is not related to the number of people infected. Thus, in other regions in China, regardless density in populations, COVID-19 have not had a major outbreak. This is supported by the finding that the correlation coefficient measuring the relation between population density and population infection in the other regions in China is 0.037.

We realized that, due to the back and forth revision and correction of the data by the official sources, it is not possible that all the data are error-free. However, these data as a whole are reliable.

In summary, our data show that the extent and speed of a large outbreak under the same conditions are directly affected by the population density, which is of great significance to the government’s decision-making on social distancing and travel restrictions when responding to the outbreak.

## Data Availability

Data are all available from public websites.

## Acknowledgments

This work was partially supported by funding from merit grant I01 BX000671 to WG from the Department of Veterans Affairs and the Veterans Administration Medical Center in Memphis, TN, USA, and grant 90DDUC0058 to CG from U.S. Department of Health and Human Services, Administration for Community Living.

## Author Contributions

Conceived and designed the experiments: HC, WG, YJ, LA, HC, DS. Performed data searching and collection: TS, HY, LY, WG. Analyzed the data: TS, HY, LY LM, LL, WG. Contributed analysis tools: WG, CH, HC, DS. Wrote the manuscript: TS, HY, LY, LA, SD, CG, SH, WG. Revise and approve and manuscript: All authors.

## Competing interests

The authors declare no competing financial interests.

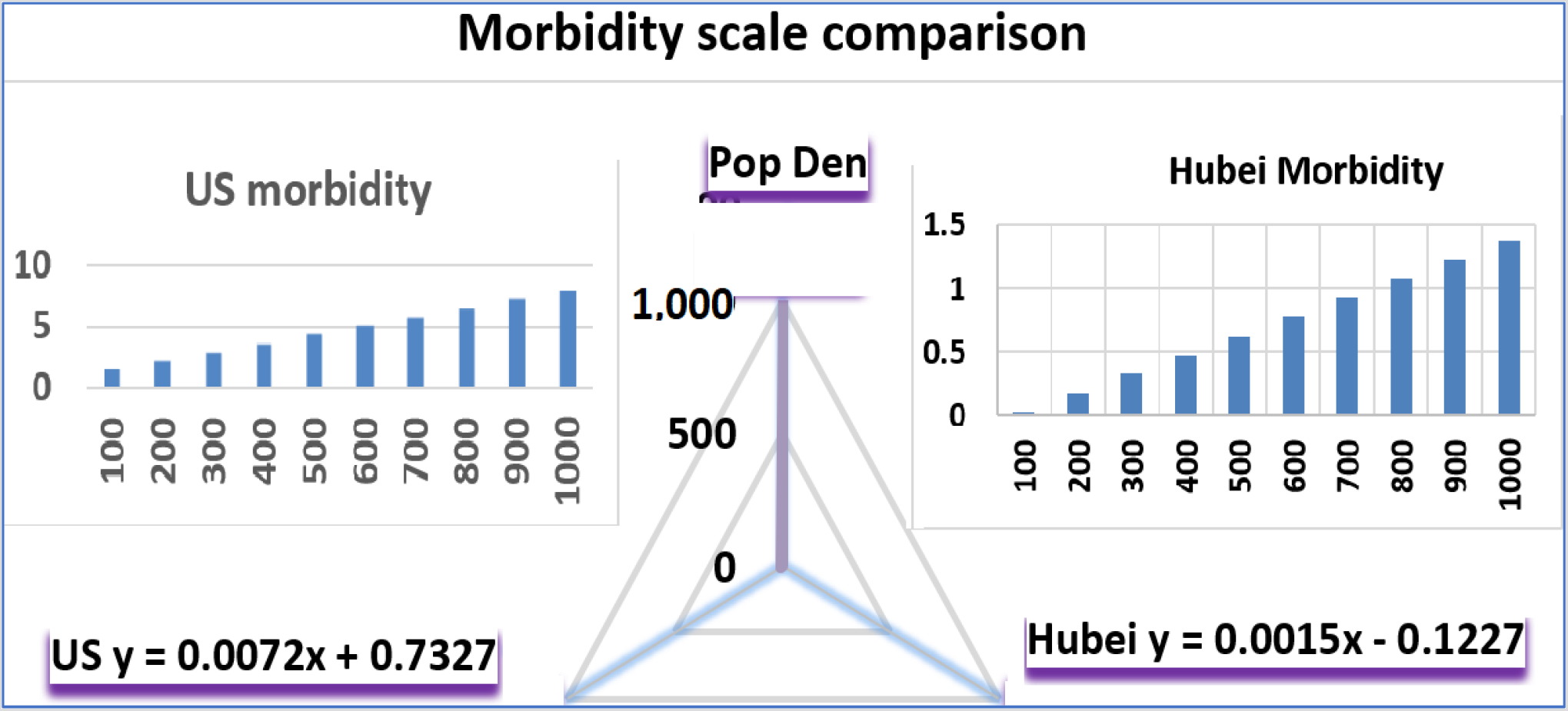

## Notes

### Competing Interest Statement

The authors have declared no competing interest.

### Author Declarations

N/A. not a clinical trial

